# Development and external validation of a logistic regression derived formula based on repeated routine hematological measurements predicting survival of hospitalized Covid-19 patients

**DOI:** 10.1101/2020.12.20.20248563

**Authors:** Stefan Heber, David Pereyra, Waltraud C. Schrottmaier, Kerstin Kammerer, Jonas Santol, Erich Pawelka, Markus Hana, Alexander Scholz, Markus Liu, Agnes Hell, Klara Heiplik, Benno Lickefett, Sebastian Havervall, Marianna T. Traugott, Matthias Neuböck, Christian Schörgenhofer, Tamara Seitz, Christa Firbas, Mario Karolyi, Günter Weiss, Bernd Jilma, Charlotte Thålin, Rosa Bellmann-Weiler, Helmut J.F. Salzer, Michael J.M. Fischer, Alexander Zoufaly, Alice Assinger

**Author notes:** Corresponding authors: Assinger A., 0043-1-40160-31405, Heber S., 0043-1-40160-31425.

## Abstract

**Background:** The Covid-19 pandemic has become a global public health crisis and providing optimal patient care while preventing a collapse of the health care system is a principal objective worldwide.

**Objective:** To develop and validate a prognostic model based on routine hematological parameters to predict uncomplicated disease progression to support the decision for an earlier discharge.

**Design:** Development and refinement of a multivariable logistic regression model with subsequent external validation. The time course of several hematological variables until four days after admission were used as predictors. Variables were first selected based on subject matter knowledge; their number was further reduced using likelihood ratio-based backward elimination in random bootstrap samples.

**Setting:** Model development based on three Austrian hospitals, validation cohorts from two Austrian and one Swedish hospital.

**Participants:** Model development based on 363 survivors and 78 non-survivors of Covid-19 hospitalized in Austria. External validation based on 492 survivors and 61 non-survivors hospitalized in Austria and Sweden.

**Outcome:** In-hospital death.

**Main Results:** The final model includes age, fever upon admission, parameters derived from C-reactive protein (CRP) concentration, platelet count and creatinine concentration, approximating their baseline values (CRP, creatinine) and change over time (CRP, platelet count). In Austrian validation cohorts both discrimination and calibration of this model were good, with c indices of 0.93 (95% CI 0.90 - 0.96) in a cohort from Vienna and 0.93 (0.88 - 0.98) in one from Linz. The model performance seems independent of how long symptoms persisted before admission. In a small Swedish validation cohort, the model performance was poorer (p = 0.008) compared with Austrian cohorts with a c index of 0.77 (0.67 - 0.88), potentially due to substantial differences in patient demographics and clinical routine.

**Conclusions:** Here we describe a formula, requiring only variables routinely acquired in hospitals, which allows to estimate death probabilities of hospitalized patients with Covid-19. The model could be used as a decision support for earlier discharge of low-risk patients to reduce the burden on the health care system. The model could further be used to monitor whether patients should be admitted to hospital in countries with health care systems with emphasis on outpatient care (e.g. Sweden).

## Introduction

### Background

The Covid-19 pandemic evokes a complex global public health crisis with clinical, organizational and system-wide challenges. As we are currently experiencing the second or even third wave of the Covid-19 pandemic in Europe as well as other parts of the world, the health care system in many countries is at risk to collapse. To date no reliable biomarker exists to predict patient outcome and severity of disease progression. Therefore, care providers are often making decisions based on individual experience or non-validated biomarkers. Given the intensive workload of health care providers worldwide, a reliable, easily accessible prognostic tool would be beneficial.

Although a plethora of prognostic models for Covid-19 were quickly published at the beginning of the pandemic to support medical decision making at a time when they were urgently needed, a large consortium including clinical scientists, epidemiologists, biologists, and statisticians, came to the conclusion that ‘the proposed models are poorly reported, at high risk of bias, and their reported performance is probably optimistic’ (Wynants et al., 2020). Based on that, the authors did ‘not recommend any of these reported prediction models for use in current practice’, while there remains an urgent need to develop more rigorous prediction models and validate promising ones. The authors recommended building on previous literature and expert opinion to select predictors, rather than selecting predictors in a purely data driven way. Promising candidates include age, body temperature, sex, blood pressure, creatinine, basophils, neutrophils, lymphocytes, alanine transaminase, albumin, platelets, eosinophils, calcium, bilirubin, creatinine, CRP and comorbidities, including hypertension, diabetes, cardiovascular disease, and respiratory disease.

Besides the critically ill patients who need to be treated in intensive care units, the multitude of patients being treated in general wards bind substantial resources that would be needed for patients with other diseases. This negatively impacts the treatment options of patients who do not have Covid-19, causing collateral medical damage. Specifically, the measures to avoid infection of personnel and spreading of SARS-CoV-2 throughout the hospital, as putting on and taking off protective clothing, are time consuming. Nevertheless, many of these Covid-19 cases cannot be discharged since the critical phase of Covid-19 frequently starts around 7-10 days after onset of the initial symptoms. Clearly, a tool reliably predicting the likelihood of a severe or fatal disease would be beneficial and could support the decision for an earlier discharge.

### Objectives

Hence, the first objective of this study was to develop a prognostic model with predictors selected based on pathophysiological considerations and literature. Importantly, the predictors had to be easily accessible routine parameters to avoid obstacles to clinical application. The second objective was to validate the model in independent cohorts and to provide a publicly available online calculator and a formula to facilitate decision making during the current pandemic.

## Methods

### Source of the data

We conducted an observational cohort study to develop and validate a prognostic model to predict in-hospital mortality of patients with Covid-19. For this purpose, only data collected in clinical routine were used. There were no additional measurements for study purposes. Within all cohorts, data of all consecutive patients were accessed. For model development cohorts of three Austrian hospitals were used: the Clinic Favoriten (former Kaiser Franz Josef Hospital) in Vienna, the Johannes Kepler University Clinic in Linz and the Medical University of Innsbruck (cohorts 1-3). The model was validated in additional consecutive patients treated in the Clinic Favoriten and other patients treated in the Johannes Kepler University Clinic in Linz as well as in the Danderyd Hospital in Stockholm, Sweden (cohorts 4-6).

### Participants

For model development 417 Covid-19 patients from three different large-scale medical centers were included, validation is based on 405 patients from two medical centers in Austria and one in Sweden (Fig. 1). SARS-CoV-2 positivity was determined from nasopharyngeal or oropharyngeal swabs via real-time polymerase chain reaction (qPCR) according to the Charité protocol (Corman et al., 2020). Routine laboratory analyses were performed within clinical routine. Additional blood withdrawals were performed if clinically indicated.

**Figure 1:**
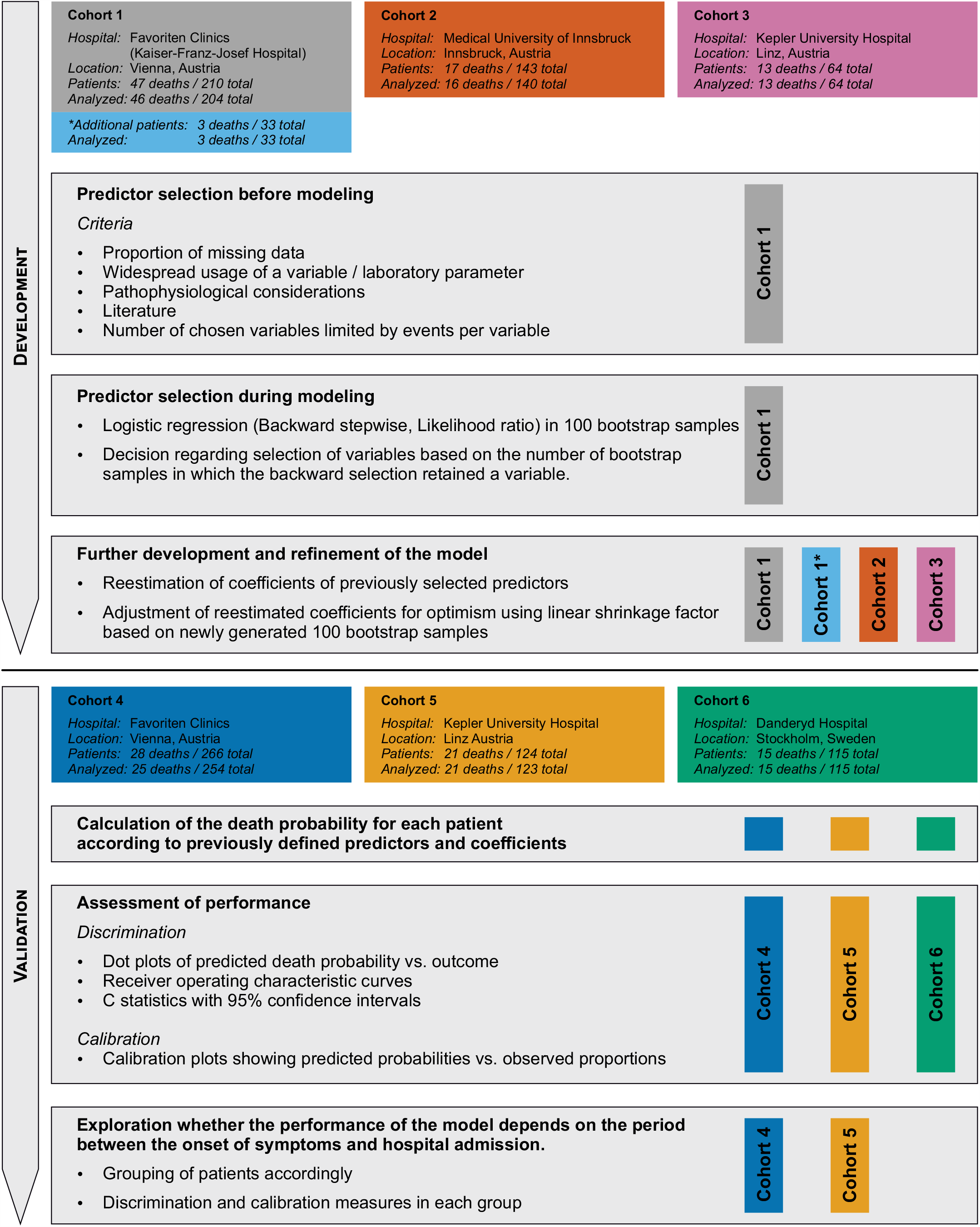
Overview of cohorts used for model development and validation.

All patients had available outcome data at time of analysis. Of note, the recovery of data at the Clinic Favoriten in Vienna is part of the ACOVACT study (ClinicalTrials.gov NCT04351724) approved by the local ethics committee (EK1315/2020), which aims to compare the effect of different antiviral and adjunctive treatments on outcome of hospitalized Covid-19 patients. This study was further approved by the ethics committee of the Innsbruck Medical University (ID of ethical vote: 1167/2020), the ethics committee of the Kepler University Clinics (1085/2020) and the Stockholm Ethical Review Board (COMMUNITY study dnr 2020-01653).

### Outcome

The predicted outcome is death from any cause during the hospital stay. There was no loss to follow up as patients were either discharged or died.

### Predictors

As the aim was to build a prognostic model that has widespread applicability, only routinely measured variables were considered, such as measurements of the blood count and other hematological parameters. Thus, the methods of measurement are the ones usually used in the respective hospital.

The first selection of predictors from all available variables was based on graphical inspection of data, providing information on the time course of variables and the proportion of missing data. Further selection of possibly useful predictors considered pathophysiological processes, the published literature, and had a special focus on the reported Covid-19-associated coagulopathy. The graphical exploration showed that although some potential predictors are not changed to a relevant extent at the time of admission, they develop considerably differently between survivors and non-survivors during hospital stay. Therefore, we included the variable’s time course in the prognostic model. Since blood samples were often taken only every two days from admission, and as data from two days seemed too short for a prognosis, the pragmatic decision was made to use the data from day 0 to 4 (i.e., 5 calendar days) after admission for prognosis. Data processing is described in detail in the statistical methods section.

### Blinding

The individuals accessing the medical records to extract variables were not blinded to the outcome.

### Sample size

A formal sample size calculation was not performed. The cohort of hospitalized patients suffering from Covid-19 continues to grow. For the development of the model, we roughly oriented ourselves on 10 events per variable, considering that this rule of thumb might be an oversimplification of the problem regarding an adequate sample size in logistic regression (Ogundimu et al., 2016; Vittinghoff and McCulloch, 2007). For validation, we also aimed for this number in each validation cohort. However, we had to consider that a model that can potentially reduce the burden on health systems should be available to the general public rather sooner than later.

### Missing data

There were no missing data regarding outcome. Regarding missing data of predictors, one has to discriminate two types in the context of our prognostic model. First, as the time course of variables over the first four days was processed by linear regression within each patient into the predictors used in the logistic regression model, the first type of missing data is missing data points within the four days after admission within a patient. As the frequency and exact time points of blood samplings within patients naturally differ between patients and clinics, we decided to fit a linear regression line if any data on this variable were available. This might, on the one hand, reduce the regression line’s correspondence of the underlying biological process, on the other hand allow application of the model even if very few blood samples are available within the first four days. The second type of missing data is missingness of the regression coefficients within a patient, if no blood samples were available, or if a certain analyte in blood samples was not determined. The proportion of patients with missing linear regression-based predictors used for logistic regression was considered low enough to exclude all patients from the analysis with such missing data. The number of patients with missing data on predictors is given in the results section with each result and in Fig. 1.

### Statistical analysis methods

#### Processing of measured values in blood samples to the predictors used in logistic regression models

Inspection of the raw data, as shown exemplary for platelet count and CRP (Fig. 2A, B), suggested that both values measured in blood samples taken upon admission and those taken at later time points contain information regarding survival. Whereas the platelet count appeared to increase throughout the hospital stay in survivors, this was not observed in patients who died. Conversely, CRP levels tended to return to baseline in survivors but increased in non-survivors. To make use of these different time courses in a prognostic model, a linear regression approach was used in each patient and for each potential predictor separately. A linear regression line was fitted through the data acquired within days 0 to 4 of hospitalization. The resulting intercept and slope were considered as potential predictors in logistic regression models. The intercept roughly reflects the value at admission. We also considered using the baseline value on admission instead of the intercept as predictor in logistic regression. However, this would have rendered the prognostic model unusable for patients without blood sampling upon admission. In contrast, the intercept as an estimate of the baseline value can also be used when, e.g., only data of day two and four are available. The slope on the other hand approximates the change over time. An example of such a linear regression within a patient’s platelet count data is shown in Fig. 2C.

**Figure 2:**
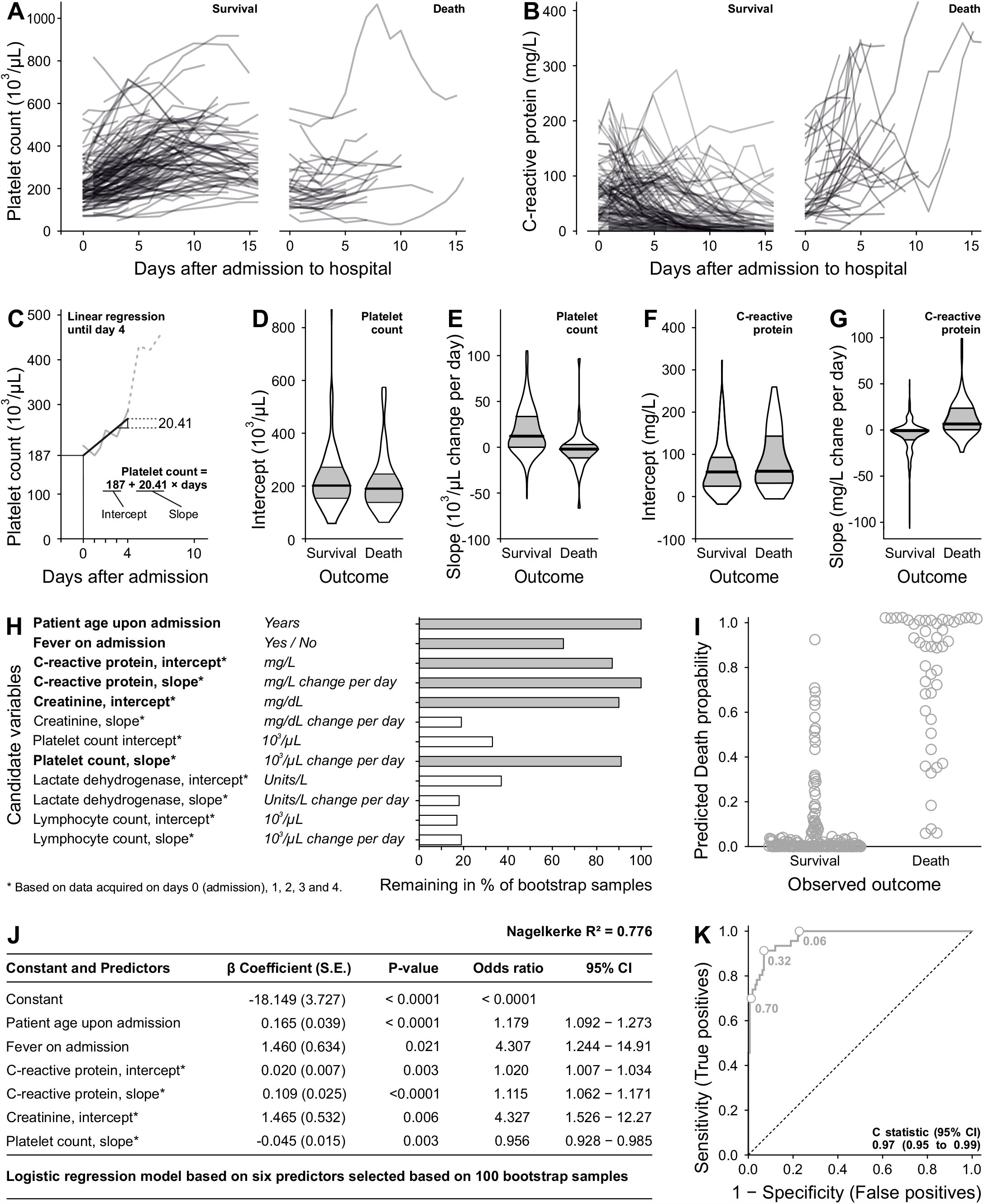
Model development in cohort 1 from Vienna. **A)** Changes of platelet count throughout the hospital stay in patients who survived and subsequently were discharged and those who died within the hospital. **B)** Analogous changes of CRP levels. **C)** Exemplary calculation of intercept and slope using linear regression. A linear regression line was fitted to all data points available from admission to day 4 and is visualized for the platelet count. The intercept approximates the value upon admission, even in case this specific value is not available in a patient. The slope reflects the change of the respective variable per day. Such a linear regression was fitted to every considered variable of every patient separately. **D)** Distribution of platelet count intercepts. Horizontal lines indicate medians, 25th and 75th percentiles. **E-G)** Distribution of platelet count slopes (E), CRP intercepts (F) and CRP slopes (G). Note that the 75th percentile of CRP slopes in survivors is so close to the median that it is hidden within the horizontal black bar indicating the median **H)** List of potential variables selected based on literature and pathophysiological considerations. For each analyte from blood samplings, intercepts and slopes were calculated. The most important predictors in 100 random bootstrap samples of the original dataset were selected separately within each bootstrap sample using logistic regression with backward stepwise selection, whereby removal testing was based on the probability of the likelihood-ratio statistic. The horizontal bars indicate the percentage of bootstrap samples in which the respective variable was retained in the final model after the stepwise procedure. Based on that, it was decided to use the variables in bold face with filled corresponding bars in the final model. **I)** A logistic regression model with regression coefficients listed in J was estimated using the original dataset and the previously selected predictors. The resulting estimated probabilities for death are plotted against the observed outcome. **J)** Regression coefficients of the model in the original dataset of Cohort 1. Nagelkerke’s R^2^ is given as a measure of effect size and represents a version of the coefficient of determination (R^2^) for logistic regression. The odds ratios result from the constant e raised to the power of the respective coefficient. The odds ratio of age, for instance, can be interpreted that each additional year of age raises the odds of death by a factor of 1.179. **K)** Corresponding ROC curve. The c statistic indicates the probability that when two random individuals are selected with different outcomes, the model assigns a higher predicted death probability to the subject who died. This probability equals the area under the ROC-curve. To illustrate which cut-off values of predicted probabilities for the discrimination between survival and death result in which combination of true and false positive rates, some exemplary cutoff values are plotted on the curve. Assuming a fatal outcome for every patient above a predicted death probability of 0.32, for example, results in a sensitivity of 91% and a false positive rate of 7%, i.e., a specificity of 93%.

#### Handling of predictors

To avoid a loss of information, all continuous potential predictors were used as such, i.e., were not categorized. Although it seems likely that body temperature in its continuous form would have contained some information regarding outcome, it was only available as dichotomous variable fever on admission, ‘Yes’ was coded as 1, ‘No’ as 0. The distributions of the continuous predictors platelet count intercept, platelet count slope, CRP intercept and CRP slope are visualized in Fig. 2D-G separately for survivors and deceased.

#### Type of model, variable selection, model building procedures

To estimate death probability, multivariable binary logistic regression models were used. A subject matter knowledge-based pre-selection of potential predictors was determined before modeling and is listed in Fig. 2H. We aimed to further reduce the number of variables during modeling for two reasons. First, we wanted the final model to be as simple as possible for a high degree of clinical applicability. Second, we aimed to have a number of events per variable that at least approximates ten. A bootstrap approach was chosen for this purpose, whereby 100 random bootstrap samples were generated from data of cohort 1 at first. Following this, the pre-selection of predictors was entered and subsequently removed stepwise backward in logistic regression models built based on each bootstrap sample separately. Which variables remained in each model was based on the significance of the change in the log-likelihood upon removal of a variable. The percentage of bootstrap samples in which a candidate variable was not removed is plotted in Fig. 2H. Those variables that were retained most often in the model were chosen. Although fever on admission was retained only in approximately two thirds of bootstrap samples, it seemed likely that fever on admission contains information regarding the outcome, wherefore it was retained in the model. Univariable predictor outcome analyses were not performed. Due to the limited number of events at this stage, interactions were not tested to avoid overfitting.

After the final selection of the predictors was determined, we decided to further develop the model before external validation. To this end we added patient data to the already existing dataset. As most of the additional patient data were from different cities in Austria than the original one, we expected that generalizability of the model would be improved. Based on the combination of the original data and the new data all regression coefficients were re-estimated. The resulting model was adjusted for optimism using a linear shrinkage factor (Steyerberg et al., 2001). In short, 100 random bootstrap samples with the same sample size as the original dataset were generated from the available data (i.e., cohorts 1-3). Within each bootstrap sample, a logistic regression model was fitted using the previously determined predictors. This resulted in different beta coefficients for each predictor in each bootstrap sample. Using the coefficients of these 100 logistic regression models, the prognostic indices were calculated for each patient in the original sample. The prognostic index is the linear combination of the regression coefficients as estimated in the bootstrap sample with the values of the covariables (e.g., age) in the original sample. Next, the prognostic index derived from each bootstrap sample was used as a single covariable in logistic regression analyses in the original dataset. The beta coefficient of the single covariable ‘prognostic index’ reflects the slope of the prognostic index. For each bootstrap sample, a slightly different slope occurred. The mean of these slopes, usually below 1, reflects that the regression coefficients are too extreme for prognostic purposes and was used as linear shrinkage factor, i.e., the coefficients were multiplied by this factor. The result was the final prognostic formula subsequently validated externally.

External validation was performed by calculating the death probabilities of additional patients using the established formula and calculating performance measures. Thereby, patients that were treated at the Favoriten Clinics in Vienna (Cohort 4) and at the Kepler University Hospital, Linz (Cohort 5) served for validation purposes in the sense of temporal validation. We also validated the model in a cohort from the Danderyd hospital in Stockholm, Sweden (Cohort 6).

In addition to plotting the predicted death probabilities of each patient against the outcome, we used the area under the receiver operating characteristic (ROC) curve (equivalent to the c statistic) as measure for discrimination. For assessment of calibration, we plotted the observed risks on the y-axis versus the predicted risk on the x-axis. Specifically, we first generated five groups of patients. The groups were defined by predicted death probabilities: i) 0 to ≤ 0.2, ii) > 0.2 to ≤ 0.4, iii) > 0.4 to ≤ 0.6, iv) > 0.6 ≤ 0.8, v) > 0.8. Within each group, the mean of all probabilities was calculated, which served as an x-axis coordinate for the plotted symbol. The y-axis value was determined by the proportion of deaths in the respective group. To visualize the uncertainty regarding these probabilities, 95% confidence intervals were calculated as described (Brown et al., 2001). Due to the relatively low number of patients, especially those with higher predicted death probabilities, five groups were chosen instead of the usual ten groups. The performance of the model in the two Austrian validation cohorts was compared with the performance of the same model in the Swedish cohort by the probability P of the z-value corresponding to the AUC difference calculated as follows:

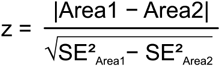

#### Risk groups

Risk groups were not generated, as this generally might not be in the best interest of patients (Steyerberg, 2009). Additionally, groups do not seem meaningful in the context of how our model could be used clinically: To assign insufficient resources, the patients with the lowest death probability could be discharged.

#### Differences between development and validation

As this study encompasses both development and validation, differences in definitions and variables including their coding are not of concern. However, in the Swedish cohort, outcome of in-hospital mortality could in principle be affected by differences in admission and discharge practices. This might affect the proportion of Covid-19 caused deaths occurring in hospital and thus counting as an event.

## Results

### Participants

An overview of the cohorts including the number of survivors and non-survivors is shown in Fig. 1. Patient characteristics are listed in Tables 1 and 2.

**Table 1.**
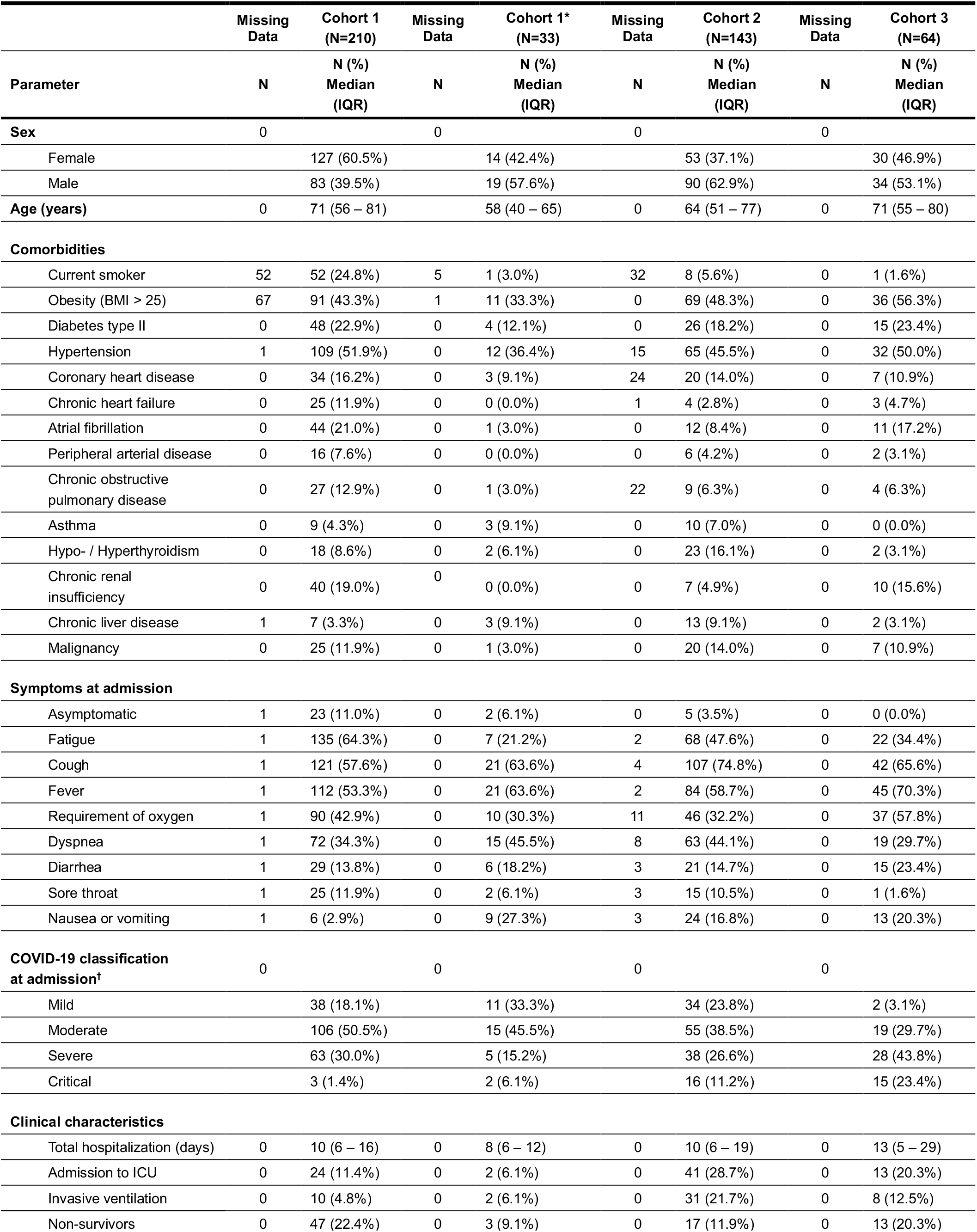
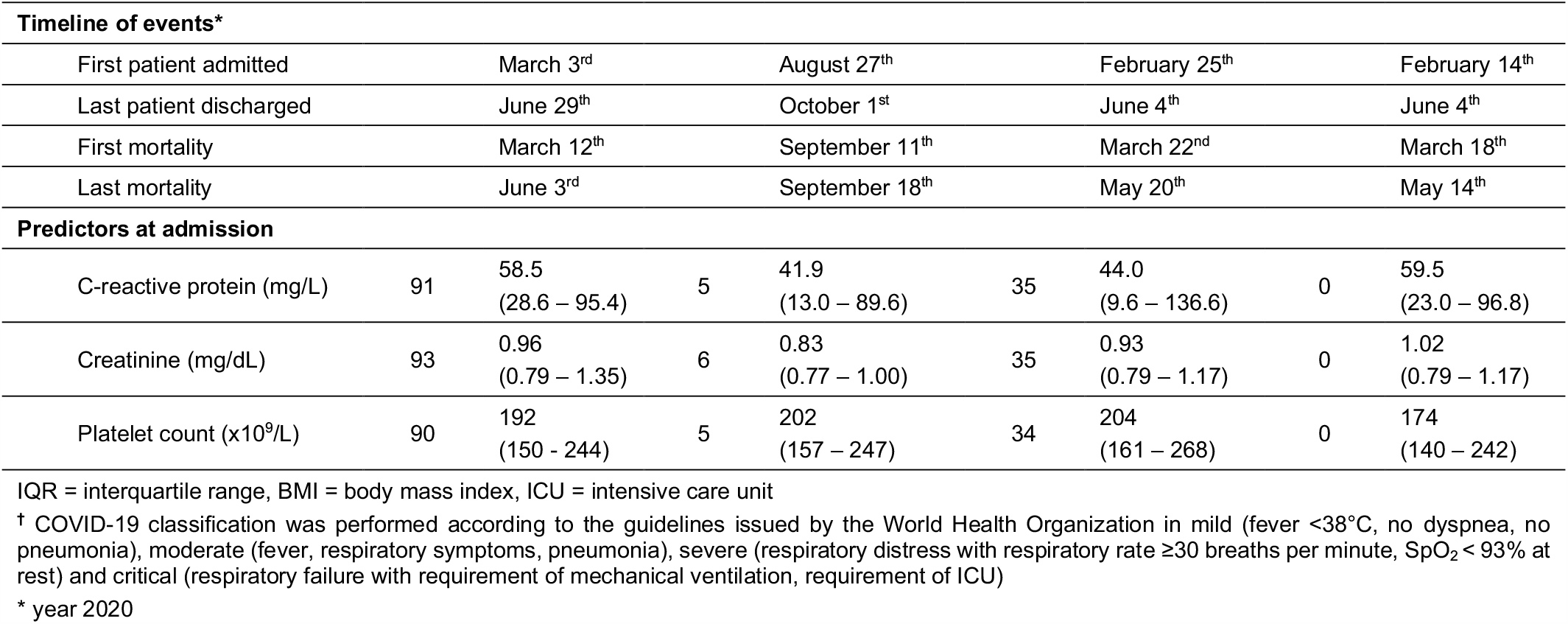
Patient demographics for Development Cohorts.

**Table 2.** Patient characteristics of Validation Cohorts.

|  | Missing Data | Cohort 1-3 (N=450) | Missing Data | Cohort 4 (N=266) | Missing Data | Cohort 5 (N=124) | Missing Data | Cohort 6 (N=115) |
| --- | --- | --- | --- | --- | --- | --- | --- | --- |
| Parameter | N | N (%)<br>Median<br>(IQR) | N | N (%)<br>Median<br>(IQR) | N | N (%)<br>Median<br>(IQR) | N | N (%)<br>Median<br>(IQR) |
| Age (years) | 0 | 67<br>(52 - 79) | 0 | 56<br>(44 - 70)*** | 0 | 71<br>(53 - 83) | 0 | 61<br>(50 - 69)*** |
| Fever at admission | 3 | 262<br>(58.2%) | 3 | 148 (55.6%)*** | 0 | 70<br>(56.5%) | 0 | 89<br>(77.4%)*** |
| Non-survivors | 0 | 80<br>(17.8%) | 0 | 28<br>(10.5%)** | 0 | 21<br>(16.9%) | 0 | 16<br>(13.9%) |
| **Predictors at admission** |  |  |  |  |  |  |  |  |
| C-reactive protein (mg/L) | 131 | 53.0<br>(22.0 - 98.0) | 48 | 31.1<br>(10.0 - 78.2)*** | 13 | 33.0<br>(8.0 - 77.0)* | 2 | 95.0<br>(45.0 - 166.5)*** |
| Creatinine (mg/dL) | 134 | 0.95<br>(0.79 - 1.20) | 47 | 0.86<br>(0.72 - 1.03)*** | 12 | 1.07<br>(0.90 - 1.31)** | 6 | 0.94<br>(0.71 - 1.12) |
| Platelet count ($\times 10^9/\text{L}$) | 129 | 192<br>(152 - 254) | 46 | 195<br>(155 - 243) | 9 | 204<br>(158 - 246) | 3 | 204<br>(157 - 288) |

IQR = interquartile range
\* $p < 0.05$ , \*\* $p < 0.01$ , \*\*\* $p < 0.001$ vs Cohort 1-3

### Model development

#### Predictor selection during modeling

After the subject matter-based pre-selection of predictors, their selection was further narrowed down using a bootstrap approach with data of cohort 1. Of 210 patients in this cohort, 6 were not included in the analysis since at least one predictor had missing data. One of these 6 patients died, leaving 46 deaths to be included in the analysis. The extent of the loss of information due to these 6 patients seemed negligible. Therefore, multiple imputation was abstained from and only complete cases were analyzed. Results (Fig. 2H) suggested that the following predictors should be retained in the model: 1) patient age in years, 2) fever on admission (binary, with fever being defined as a body temperature > 38°C), 3) the intercept of the linear regression line through CRP levels over time, approximating the level upon admission, 4) the slope of this regression line, reflecting the change per day, 5) the intercept of the regression line through creatinine levels and 6) the slope of the regression line through the platelet count. The model including the selected predictors fitted on the original (non-bootstrapped) data led to the predicted death probabilities shown in Fig. 2I. The corresponding regression coefficients and ROC curve can be found in Fig. 2J and K. The apparent performance measured by the c statistic was 0.97 (95% CI 0.95 - 0.99), Nagelkerke’s pseudo R^2^ measure of 0.776 indicated a large effect size.

#### Further development and refinement of the model

The initial model based on cohort 1 was updated by adding 33 more patients of the same hospital (referred to as additional patients of cohort 1), cohort 2 comprising 143 patients and cohort 3 consisting of 64 patients. Of the total of 450 patients, 9 had missing data, leaving 363 survivors and 78 non-survivors. Reestimation of all regression coefficients led to a slightly reduced effect size indicated by a Nagelkerke’s R^2^ of 0.597 (Fig. 3A). The predicted probabilities of all cohorts are plotted in Fig. 3B, to allow inspection of predicted probabilities in each cohort they are also plotted separately by cohort (Fig. 3C-E). The ROC curve corresponding to the predicted death probabilities in Fig. 3B is plotted in Fig. 3F. The adjustment of the regression coefficients for optimism using a linear shrinkage factor (Fig. 3G) resulted in the shrunken coefficients (Fig. 3H).

**Figure 3:**
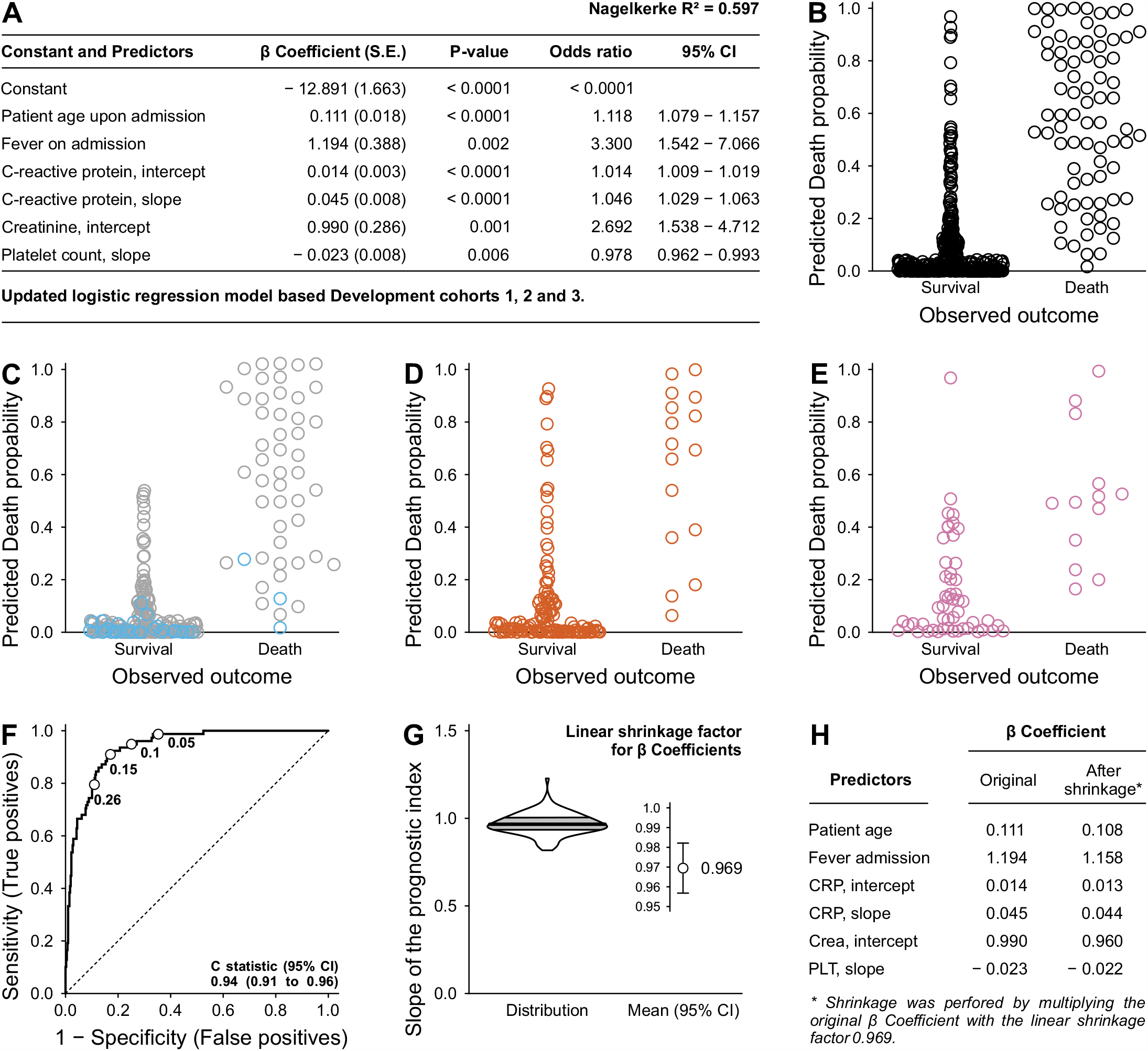
Further development of the model into its final version. The data of additional patients were combined with the data used to select predictors and the resulting total dataset was used to reestimate all regression coefficients. **A)** Regression coefficients, odds ratios, and effect size of the updated model. **B)** Predicted death probabilities plotted against observed outcome. **C-E)** Data shown in B plotted separately for cohorts to enable assessment of the model’s performance in different cohorts. C) Predicted death probabilities of patients used for predictor selection in grey, and some additional patients from the same hospital (Clinic Favoriten in Vienna) in sky blue. D) Predicted death probabilities of patients treated at the Department of Internal Medicine II, Medical University of Innsbruck, Innsbruck, Austria. E) Corresponding death probabilities for patients treated at Department of Pulmonology, Kepler University Hospital and Johannes Kepler University, Linz, Austria. **F)** ROC curve corresponding to death probabilities plotted in B. **G)** To adjust the regression coefficients for optimism, a shrinkage approach with a linear shrinkage factor was applied (Steyerberg et al., 2001). In short, the regression coefficients were estimated in each bootstrap sample. Next, the prognostic index, i.e., the result of the linear combination of the regression coefficients, was determined in the original dataset for each patient based on the coefficients of each bootstrap sample. This resulted in 100 prognostic indices for each patient in the original data, corresponding to the 100 bootstrap samples. Subsequently, the prognostic indices for each patient were used as a single covariable in a logistic regression using the original dataset. The coefficient of the covariable, i.e., the slope, naturally differs between prognostic indices derived from different bootstrap samples. The violin plot shows the distribution of the slopes based on the 100 bootstrap samples, the circle with the error bar represents the mean. The mean slope less than 1 indicates that the original coefficients are too extreme for predictive purposes, but with a mean slope of 0.969 there was only a slight overestimation. **H)** Shrinkage of the original coefficients according to the mean slopes, i.e., the linear shrinkage factor.

#### Model specification and performance

The explicit formula of the model is given in Fig. 4A. To calculate a patient’s risk one first needs to fit linear regression lines as described in the methods section to obtain intercepts and slopes for CRP, creatinine, and platelet count. Following this, the patient’s age in years, the information whether fever was present upon admission (1 for ‘Yes’ and 0 for ‘No’), intercepts (abbreviated with ic) and slopes (sl) need to be entered in the formula. The result is the predicted death probability. Alternatively, the values in their original form (e.g. the day after admission with the platelet count in thousands / µl measured at that day) can be entered into the online calculator.

**Figure 4:**
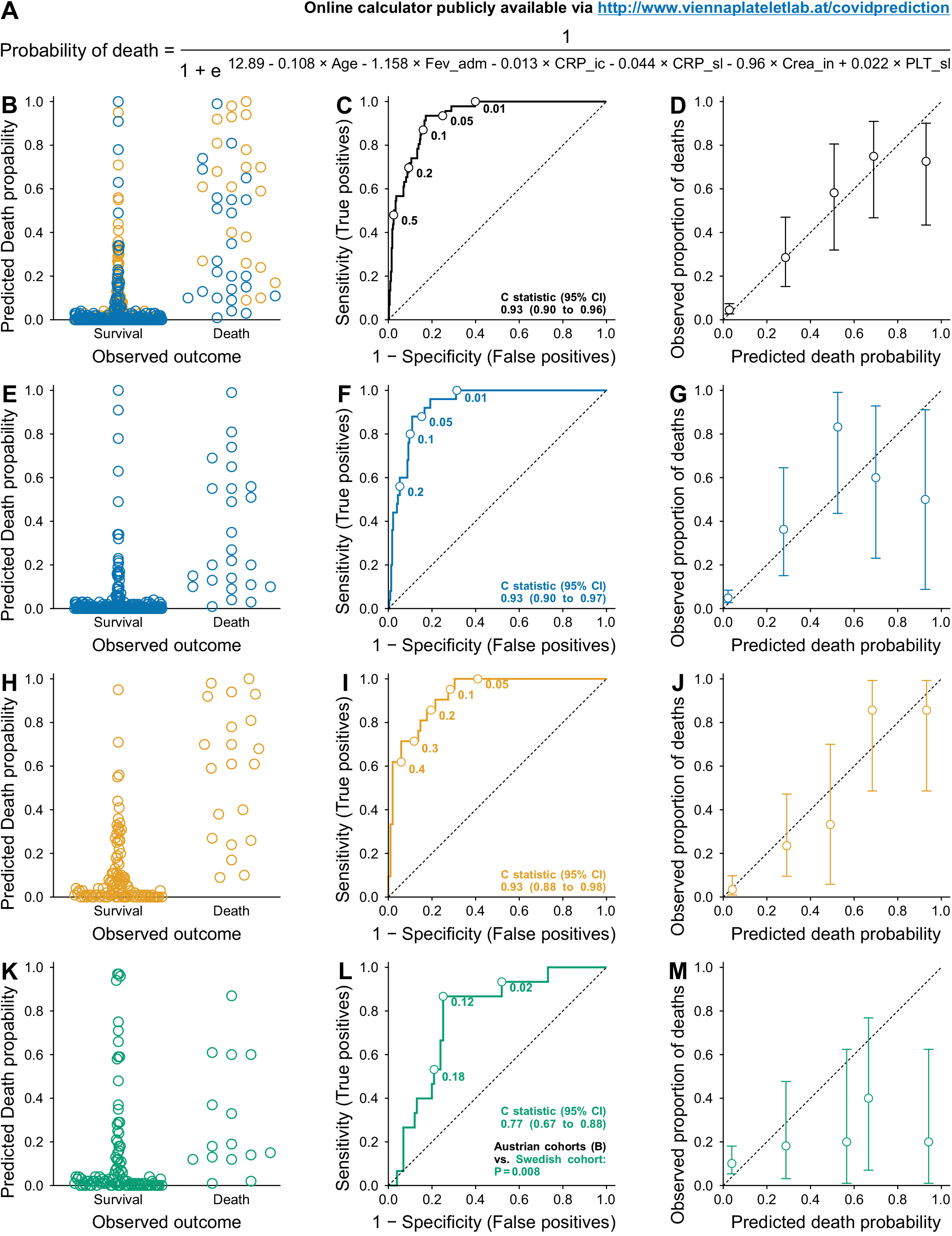
External validation of the prognostic model in cohorts from Austria and Sweden. **A)** Final prognostic model subsequently validated in independent patient cohorts with link to the online calculator. **B)** Predicted death probabilities of survivors and non-survivors. Blue symbols indicate patients from the Clinic Favoriten in Vienna, Austria, orange patients from the Kepler University Hospital in Linz, Austria. **C)** Corresponding ROC curve with exemplary cut-off values and c statistic. **D)** Calibration plot corresponding to data shown in A and B. Ideally, all symbols should lie on the dotted diagonal line of identity, indicating that the predicted death probabilities correspond exactly to the observed ones. **E-G)** Corresponding plots for the patients from Kaiser Franz Josef hospital. **H-J)** Corresponding plots from the Kepler University Hospital in Linz, Austria. **K-M)** Data from Danderyd Hospital, Stockholm, Sweden. Corresponding plots show that the model performed significantly and relevantly worse compared to the Austrian cohorts.

The probabilities calculated in this way are presented in Fig.4B separated by outcome, but pooled for the two Austrian validation cohorts, with the corresponding ROC curve in Fig. 4C. The c statistic of 0.93 (95% CI 0.90 - 0.96) indicates that the formula will assign a higher predicted death probability to a random subject that died compared to a random subject that survived in 93% of cases. The calibration plot (Fig. 4D) serves to compare the predicted probabilities over the whole range of probabilities with the actual outcome. Ideally, the symbols are exactly positioned on the diagonal dotted line of identity, which indicates identity of the predicted death probability with the observed proportion of deaths. The 95% confidence intervals, being a measure of uncertainty of the observed proportions, are wider towards higher death probabilities as there were less patients with a high probability to die than there were patients with a lower probability. Overall, the calibration plot shows a clinically useful calibration, with relatively precise prediction of death in patients with a low death probability, which is relevant for the model’s intended use, and a slight overestimation of death probabilities in patients with a high proportion of deaths. Respective separate plots for cohorts 4 and 5 (Fig. 4 E-J) show discrimination and calibration measures for cohort 4 and 5 separately, suggesting similar performance in both cohorts.

However, the model performed significantly worse in the Swedish validation cohort (Fig. 4K-L). Although still significant, as indicated by a 95% confidence interval of the c index not including 0.5, both discrimination and especially calibration seem clinically less useful. Of note, patients from Sweden showed a longer duration between symptom onset and admission to the hospital compared to both Austrian validation cohorts (Fig. 5).

**Figure 5:**
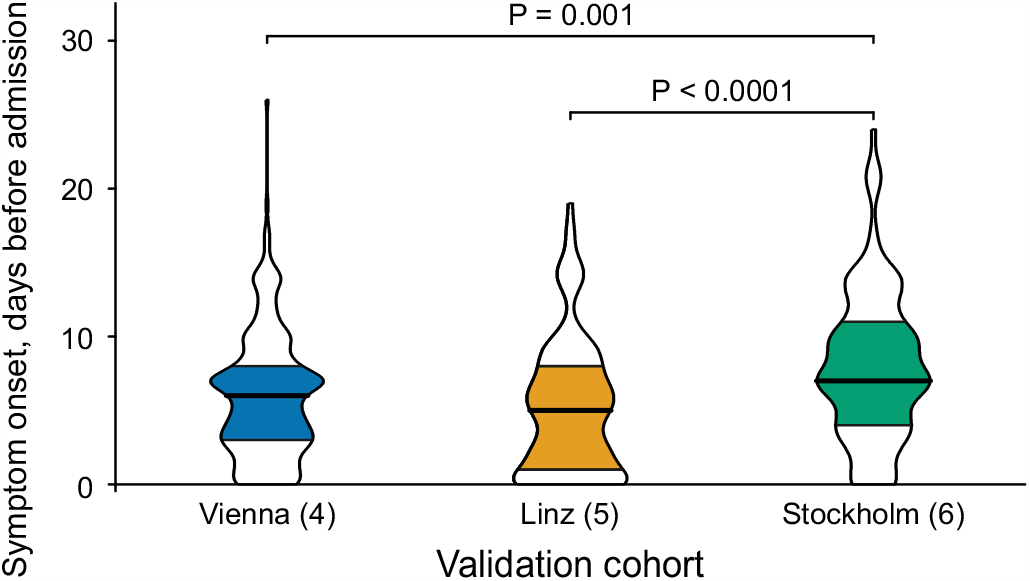
The period between symptom onset and admission to the hospital in the Austrian (Vienna, Linz) cohort is shorter than in the Swedish validation cohort. P-values are the result of Mann-Whitney tests adjusted for three pairwise tests according to Bonferroni’s method. Vienna vs. Linz P = 0.123.

#### Model performance in relation to the lag time between symptom onset and hospital admission

It is particularly challenging to assess the future disease course of Covid-19 patients who have had a more recent onset of symptoms. Therefore, it is clinically important that the model also performs well in such a patient collective. To explore this issue, we grouped all Austrian validation patients (cohorts 4 and 5) according to their self-reported symptom onset. Regarding the Swedish cohort, the event count is currently insufficient to perform an analogous meaningful subset analysis. As measured by the c indices, the model’s performance seems to be roughly stable over the range of periods between symptom onset and admission in Austrian cohorts (Fig. 6). However, due to the low number of events, especially in those patients with a long symptom duration before admission, these exploratory results need to be interpreted with great caution. Still, even in those patients where the symptom onset could not be verified, the model seems to discriminate between patients who die and those who survive.

**Figure 6:**
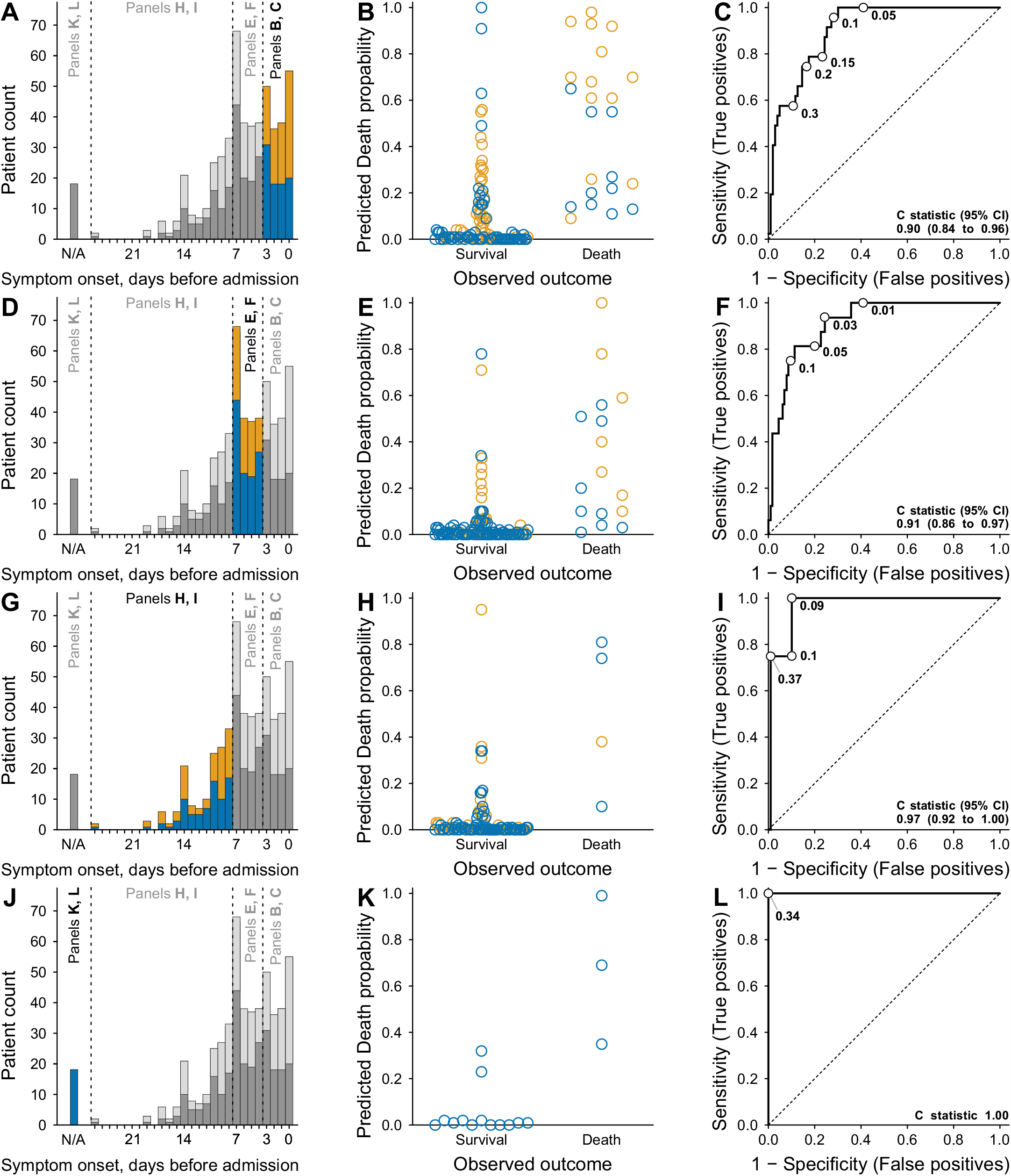
Effect of the period between the self-reported symptom onset and the admission to the hospital on the model’s performance in Austrian validation cohorts. **A)** Histogram depicting the number of patients according to the delay between the onset of symptoms and admission to the hospital. Blue indicates patients from the Clinic Favoriten in Vienna, Austria, orange patients from the Kepler University Hospital in Linz, Austria. **B)** Predicted death probabilities plotted against the observed outcome for patients whose symptoms began within three days before admission to hospital with **(C)** the corresponding ROC curve. **D-F)** Corresponding plots showing patients whose symptoms began between the fourth and seventh day before admission to hospital. **G-I)** Corresponding plots showing patients whose symptoms began more than one week before admission to hospital. **J-L)** Corresponding plots for patients whose symptom onset could not be determined, e.g., due to language barriers. I and L) The ROC curves are depicted for completeness; however, data need to be interpreted with great caution due to the low sample size and event rate in these patients.

#### Model updating

The model was not updated after validation.

## Discussion

The main result of this study is a simple formula that predicts the risk of death of hospitalized patients with Covid-19 within the period of their stay. This information could be used as additional decision support regarding discharge of clinically stable Covid-19 patients in case adequate therapy is also available at home. The formula is based on predictors routinely measured in hospitals, which allows immediate and widespread use. This is also facilitated by a publicly available online calculator.

### Limitations

The most important limitation is the uncertainty whether the model performs adequately in other geographical regions than Austria. Validation in a relatively small Swedish cohort showed a significantly worse model performance than in two Austrian cohorts. However, we have to state that there are substantial differences between the Austrian and the Swedish cohorts, which could explain the discrepancy in the performance. First, admission criteria differed between Austria and Sweden when the patients included in this analysis were hospitalized. While Swedish patients were only admitted if they needed additional oxygen, the indication for hospitalization was far more permissive in Austria. Consequently, patients in Sweden were hospitalized at a later stage of the disease, as evidenced by the significantly higher number of days with symptoms before admission in the Swedish cohort compared to the Austrian ones. Further, the peak of the first wave of the pandemic hit Sweden far stronger than Austria, which also affected clinical care differently. For instance, few patients from nursing homes were admitted to Swedish hospitals at that time, explaining why the Austrian cohorts show a wider age distribution compared to the Swedish cohort. Of note, while in general mortality rates during spring were much higher in Sweden compared to Austria, the percentage of in hospital mortalities was much lower in this cohort compared to Austria (12.5% in Stockholm versus 22.4% in Vienna at the same period). This indicates that the hospitalized Swedish cohort represents only a fraction of severe Covid-19 cases, while in Austria a larger fraction of patients with severe symptoms were treated in hospital. We can therefore not completely rule out that if outpatient data would have been included the performance would have been similar in both countries. Hence, it is necessary to validate and possibly adapt the model for other regions.

The model in its original form might also be repurposed for the decision whether hospitalization should take place. For example, regular blood samplings could be done in Covid-19 positive patients in nursing homes or outpatient clinics. As soon as the model indicates a death probability substantially above zero, a patient could be admitted to hospital to provide optimal care. Importantly, validation of the model for this repurposing is warranted.

Another limitation is inextricably linked to the way this prognostic model is built. The fact that data are used from the first five calendar days (including the day of admission) of the hospital stay means that a prognosis can only be made after this period. However, based on 920 German hospitals with over 10,000 patients (Karagiannidis et al., 2020), even the less critical non-ventilated patients had a median stay of 9 days (interquartile range 5-15 days), while critical (ventilated) patients stayed much longer. Therefore, despite this limitation, we believe that earlier discharge of non-critical patients based on our model could save resources that might benefit patients in a more critical condition.

There is another caveat regarding the interpretation of the calculated death probabilities. As can be seen in the calibration plots of the validation in Austrian cohorts (Fig. 4D, G, J) in the lower left corners, the lower predicted probabilities agree well with the observed proportion of deaths. Confidence in this regard is quite high, as indicated by the relatively small error bars reflecting the 95% confidence intervals. These narrow intervals result from a large sample size of patients at low risk. Conversely, the upper right corner shows that the probabilities of death appear to be overestimated, and that the corresponding uncertainty indicated by the wide 95% confidence intervals is high. Thus, while the model might be quite accurate in case the true underlying proportion of deaths is in the range of the upper end of the confidence interval plotted, i.e., near the line of identity, it might also be that the model markedly overestimates the probabilities of death if the actual proportion of non-survivors is more likely to be at the lower end of the confidence interval. Notably, this limitation does not affect the clinical usability of the model because only the lowest probabilities would result in consequences, i.e., discharge, whereas high estimated probabilities would not have practical implications. From our point of view, it is therefore essential to convey to patients that a high predicted probability of death is associated with considerable uncertainty and must therefore under no circumstances be interpreted as a certain death verdict. On the other hand, much greater confidence can be placed in a very low estimated probability of death.

### Interpretation

#### Model performance in validation datasets compared to development data

As this study contains both development and validation of the model, differences in predictor definitions do not apply. Also, the validated model is identical to the final version of development. As could already be expected by the shrinkage factor relatively close to 1, the model performance was good in the Austrian validation cohorts originating from the same hospitals as the patient data used for development. However, the model performed not as good in a small Swedish cohort, potentially due to discrepancies outlined in the limitation section.

#### Overall interpretation

Our prognostic model is far from being the first. However, as many of the previously published ones must be assumed to be at high risk for bias (Wynants et al., 2020), we believe that our model can make a valuable contribution to relieving the burden on health systems. Compared to other well developed and validated models, e.g., the 4C mortality Score (Knight et al., 2020), ours distinguishes patients who die from those who survive better than many others, indicated by a c index well above 0.9. However, this good performance may be geographically limited. As a result, we can only recommend the use of the model in Austria before the model has been validated in other regions and, if necessary, adapted to them. In addition, we strictly adhered to the TRIPOD reporting guideline (Moons et al., 2015) and provide the whole datasets to allow our work to be scrutinized.

Another aspect that discriminates our model from others is the use of the time course of variables. Many others merely included the values at admission, which is reasonable, as information regarding prognosis should be available as early as possible. However, as our data show, many differences between survivors and non-survivors only develop over the course of a few days. While the use of this information is a major strength of our model, it could also be one of its possible drawbacks as decision making takes until day 4 of hospitalization. Clinical variables considered and finally included in our model have been included in other prognostic models for Covid-19, and are biologically plausible. The underlying pathogenesis of Covid-19 seems complex, yet four main intertwined loops (the viral, the hyperinflammatory, the non-canonical renin-angiotensin system (RAS) axis and the hypercoagulatory loop) responsible for patient deterioration have been identified. Three out of the four loops are represented in our model. The pathology starts with the viral loop and is rapidly followed by the second loop, the hyperinflammatory loop, which is represented by CRP in our model. CRP levels positively correlate with lung lesions and disease severity in Covid-19 (Manson et al., 2020; Wang, 2020). Therefore, daily monitoring CRP values in hospitalized Covid-19 patients has been suggested to facilitate risk stratification and prognostication (Sharifpour et al., 2020). This suggestion is implemented by the CRP intercept and slope and is in agreement with a recent paper demonstrating that diversion of inflammatory marker trends over time predict a fatal or good outcome of severe Covid-19 (Manson et al., 2020).

Lymphocyte counts have been suggested previously as prognostic markers as well. Although changes in leukocyte counts are clinical indicators of disease progression in Covid-19, investigating the changes in lymphocytes might be important for prediction (Fouladseresht et al., 2020). However, based on the variable selection method employing random bootstrap samples inclusion of this parameter as intercept and slope would not have led to a relevant improvement of the model.

Further, LDH is related to inflammation and cell damage and has been suggested as a risk factor for severe Covid-19 (Chen et al., 2020; Poggiali et al., 2020). However, both intercept and slope remained in less than 20% of the bootstrap samples, which indicated that this variable was less important and was therefore dropped from the model.

In addition, the third loop, the non-canonical renin-angiotensin system (RAS) axis loop was described, which is in a broader sense represented by creatinine in our model. Kidney involvement in Covid-19 is common and associated with high mortality and was described to serve as an independent risk factor for all-cause in-hospital mortality in patients with Covid-19 (Ali et al., 2020). Renal viral tropism has been reported, which is also associated with age and comorbidities as well as decreased survival (Braun et al., 2020). Data for more than 17 million people in the UK suggest that patients with chronic kidney disease are at higher risk for adverse events in Covid-19 than those with other known risk factors, including chronic heart and lung disease. (Gansevoort and Hilbrands, 2020). The fact that the slope of creatinine seemed to have less prognostic value than the intercept might reflect the importance of chronic kidney disease.

The fourth loop is the hypercoagulatory loop, which is represented by platelet count in this model. A meta-analysis of 7,613 Covid-19 patients revealed that patients with severe disease had a lower platelet count than those with non-severe disease (Jiang et al., 2020), which is in line with our data. However, not all studies have found platelet counts to be a predictor of Covid-19 mortality (Amgalan and Othman, 2020). This would also have been the case in our cohort if only the platelet count upon admission would have been considered. Accordingly, changes in platelet count were retained in the model in the form of the slope.

Undoubtedly the most important predictor of severe Covid-19 is age. A meta-analysis of 88 articles (69,762 patients) shows that age along with CRP were strong risk-factors for ICU admission and/or mortality (Katzenschlager et al., 2020). Since the incidence and severity of many diseases are positively correlated with age, this variable could implicitly include many other diseases in the model. Moreover, aging itself leads to inflammatory and immunological changes which could directly affect the response to viral infection and accelerate disease progression in Covid-19. A recent meta-analysis of twelve studies that investigated the role of age in Covid-19 indicates that the risk increased in some but not all studies when adjusting for important age-dependent risk factors as diabetes, hypertension, coronary heart disease, cerebrovascular disease, compromised immunity, previous respiratory disease, and renal disease (Romero Starke et al., 2020).

Concerning fever, a recent meta-analysis reported that fever is a predictor of adverse outcome in Covid-19 (Li et al., 2020). In line with studies on other viral infectious diseases, a study found that prolonged fever for 7 days from onset of illness is associated with adverse outcomes from Covid-19, while saddleback fever is not indicative of adverse outcome (Ng et al., 2020). In our model fever at admission was incorporated in the prediction model. The time course of body temperature would have been interesting to include, however, available records only allowed inclusion as a binary variable.

#### Implications

For the time being, the model is primarily applicable to patients hospitalized with verified Covid-19 and should support decision making on earlier discharge. Since the model performed best within the Austrian health care system, validation of the model in different regions is required to assess where it can be used in its original form and where it needs to be adapted.

The model was developed for and its use should be restricted to a specific clinical application, if not validated for other purposes. In case the number of patients with Covid-19 in the general ward exceeds numbers that can easily be handled and thus binds resources that would be urgently needed elsewhere, the attending physicians could decide to discharge those patients with the lowest model-predicted death probabilities. It is vital that the estimated probability is not the sole criterion for decision-making and that the physician should always include a further assessment of the situation. Furthermore, it should be ascertained that discharge has no relevant impact on treatments, i.e., only those patients should be discharged where an adequate treatment can be implemented on an outpatient basis or in quarantine. It is necessary to emphasize that high estimated death probabilities should not be overinterpreted, as their reliability is not as well determined as low death probabilities.

There is no general recommendation for a cutoff value, below which an earlier discharge would be justified. This cutoff depends on current strain on the health care system. In case patients need to be discharged, one could start with the ones with the lowest death probabilities. In this case, the ROC-curves provide the proportion of true and false positives for a specific cutoff.

In countries which rely on outpatient care, the model could be repurposed to decide whether patient hospitalisation is indicated to provide optimal treatment for patients requiring intensive care. Future research should include validation of the model in different geographical regions, repurposing for outpatient care and updating, as standard of care and the pandemic evolve.

## Data Availability

All original data will be made available upon request.

